# Excess mortality during the COVID-19 outbreak in Italy: a two-stage interrupted time series analysis

**DOI:** 10.1101/2020.07.22.20159632

**Authors:** Matteo Scortichini, Rochelle Schneider dos Santos, Francesca De’ Donato, Manuela De Sario, Paola Michelozzi, Marina Davoli, Pierre Masselot, Francesco Sera, Antonio Gasparrini

## Abstract

**Background:** Italy was the first country outside China to experience the impact of the COVID-19 pandemic, which resulted in a significant health burden. This study presents an analysis of the excess mortality across the 107 Italian provinces, stratified by sex, age group, and period of the outbreak.

**Methods:** The analysis was performed using a two-stage interrupted time series design using daily mortality data for the period January 2015 – May 2020. In the first stage, we performed province-level quasi-Poisson regression models, with smooth functions to define a baseline risk while accounting for trends and weather conditions and to flexibly estimate the variation in excess risk during the outbreak. Estimates were pooled in the second stage using a mixed-effects multivariate meta-analysis.

**Results:** In the period 15 February – 15 May 2020, we estimated an excess of 47,490 (95% empirical confidence intervals: 43,984 to 50,362) deaths in Italy, corresponding to an increase of 29.5% (95%eCI: 26.8 to 31.9%) from the expected mortality. The analysis indicates a strong geographical pattern, with the majority of excess deaths occurring in northern regions, where few provinces experienced up to 800% increase during the peak in late March. There were differences by sex, age, and area both in the overall impact and in its temporal distribution.

**Conclusions:** This study offers a detailed picture of excess mortality during the first months of the COVID-19 pandemic in Italy. The strong geographical and temporal patterns can be related to implementation of lockdown policies and multiple direct and indirect pathways in mortality risk.

**Key Messages:**

- This study evaluated mortality trends in Italy during the COVID-19 pandemic, reporting an excess of 47,490 (95% empirical confidence intervals: 43,984 to 50,362) deaths in the period 15 February – 15 May 2020, corresponding to an increase of 29.5% (95%eCI: 26.8 to 31.9%) from the expected mortality.
- There is a strong geographical pattern, with 71.0% of the estimated excess deaths occurring in just three northern regions (Lombardy, Veneto, and Emilia-Romagna), and few provinces showing increases in mortality up to 800% during the peak of the pandemic.
- The impact was slightly higher is men compared to women, with 24,655 and 23,125 excess deaths respectively, and varied by age, with higher mortality in the group 70-79 years old and evidence of a lower but measurable risk even in people less than 60.
- The analysis by week suggests differential trends, with more delayed impacts in women and elderly, and the risk limited to the early period in Central and Southern Italy, likely related to the implementation of lockdown policies and contributions from direct and indirect risk pathways.

## Introduction

The outbreak of the SARS-CoV-2 coronavirus originated in Wuhan, China, in December 2019, and quickly affected most of the countries worldwide in the first months of 2020. The infection causes COVID-19 disease, a severe acute respiratory syndrome that can lead to hospitalization and eventually death.^1,2^ As of 6^th^ of July 2020, almost 11.5 million cases of COVID-19 have been reported, resulting in 535,027 official deaths.^3^

Italy has been one of the worst affected countries. The first case of SARS-CoV-2 infection not originating in China was reported on the 20^th^ of February in the province of Lodi, and the disease quickly spread across the North of the country.^4^ The number of reported cases with a validated diagnostic test rose to 1,128 by the end of the month, and peaked on 21^st^ of March with 6,557 cases per day.^4^

A series of containment policies, both at national and local level, were implemented since the start of the outbreak. The Italian Government declared the quarantine of 11 municipalities in Northern Italy on the 21^st^ of February, which was then extended to the entire region of Lombardy on the 8^th^ of March, and finally to the whole country the next day. Strict lockdown measures were implemented thereafter, including social distancing, travel restrictions, closure of all non-essential businesses and industries, and stay-at-home orders.^5^ Most of the strictest measures were not released until early May.

The pandemic put enormous pressure on the health system, and it was particularly severe in Lombardy and few other selected provinces in other northern regions, were hospitals were overwhelmed. Emergency and intensive care units (ICU) focused almost exclusively on COVID-19 patients, with admissions for other causes substantially reduced, while general practice and outpatients activities were in many cases halted to prevent the transmission of the virus.^6,7^ The first fatality officially attributed to COVID-19 in Italy occurred in Veneto on the 21^st^ of February. The number of deaths then started rising, and by the 15^th^ of May 31,610 deaths related to COVID-19 were reported.^4^

Previous assessment on the health impact focused on excess in all-cause mortality,^8-10^ an indicator that offers a quantification of the overall burden of the disease, accounting for both direct and indirect effects.^11^ However, these analyses are limited to urban areas or used data aggregated by region, and they adopted relatively simple designs based on pre-post comparisons that do not account for temporal trends and variation in other risk factors such as weather.

In this contribution, we performed an analysis of the excess mortality during the first month of the SARS-CoV-2 coronavirus outbreak across the 107 Italian provinces, stratified by sex, age group, and period of the outbreak. The assessment is based on a new dataset released by ISTAT with better coverage, and it benefits from the application of a novel two-stage interrupted time series design and flexible statistical methods. This study can therefore offer a comprehensive overview of the mortality impact of the COVID-19 pandemic in Italy.

## Methods

### Data

The analysis is based on the database released by ISTAT on 18/06/2020,^12^ which provides number of daily deaths stratified by sex and age groups for 7,904 municipalities in 107 provinces and 20 regions of Italy within the period 01/01/2015 to 15/05/2020 (Table 1). The database includes complete daily counts for 2015-2019, and early-release data for the four and a half months of 2020 for 7,270 municipalities (92% of the total) for which linkage with death registration was up to date. The mortality counts were aggregated by province in sex and age-specific daily time series, and linked with daily mean temperature data at the population-weighted centroid of the province from the ERA-5 reanalysis dataset on the Copernicus climate data store,^13^ and weekly incidence of influenza cases at regional level determined by laboratory tests.^14^

**Table 1.**
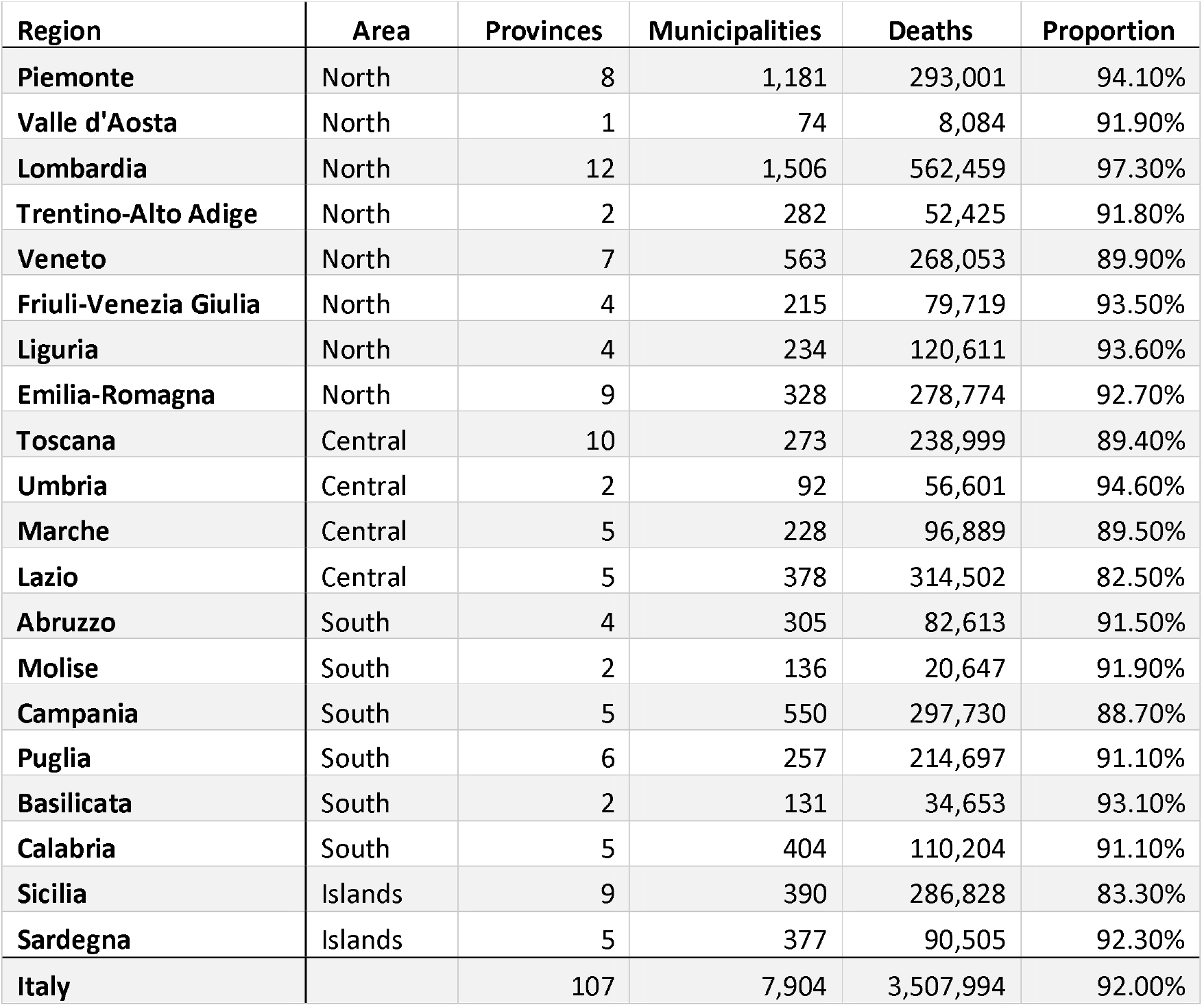
Number of provinces, municipalities, and total deaths within the study period (1^st^ January 2015 - 15^th^ May 2020) by region and area of Italy, together with the proportion of municipalities with updated data for the four months of 2020.

| Region | Area | Provinces | Municipalities | Deaths | Proportion |
| --- | --- | --- | --- | --- | --- |
| Piemonte | North | 8 | 1,181 | 293,001 | 94.10% |
| Valle d'Aosta | North | 1 | 74 | 8,084 | 91.90% |
| Lombardia | North | 12 | 1,506 | 562,459 | 97.30% |
| Trentino-Alto Adige | North | 2 | 282 | 52,425 | 91.80% |
| Veneto | North | 7 | 563 | 268,053 | 89.90% |
| Friuli-Venezia Giulia | North | 4 | 215 | 79,719 | 93.50% |
| Liguria | North | 4 | 234 | 120,611 | 93.60% |
| Emilia-Romagna | North | 9 | 328 | 278,774 | 92.70% |
| Toscana | Central | 10 | 273 | 238,999 | 89.40% |
| Umbria | Central | 2 | 92 | 56,601 | 94.60% |
| Marche | Central | 5 | 228 | 96,889 | 89.50% |
| Lazio | Central | 5 | 378 | 314,502 | 82.50% |
| Abruzzo | South | 4 | 305 | 82,613 | 91.50% |
| Molise | South | 2 | 136 | 20,647 | 91.90% |
| Campania | South | 5 | 550 | 297,730 | 88.70% |
| Puglia | South | 6 | 257 | 214,697 | 91.10% |
| Basilicata | South | 2 | 131 | 34,653 | 93.10% |
| Calabria | South | 5 | 404 | 110,204 | 91.10% |
| Sicilia | Islands | 9 | 390 | 286,828 | 83.30% |
| Sardegna | Islands | 5 | 377 | 90,505 | 92.30% |
| Italy |  | 107 | 7,904 | 3,507,994 | 92.00% |

### Two-stage analysis

The statistical analysis was performed by applying a two-stage interrupted time series model to quantify the time-varying excess risk for mortality during the COVID-19 outbreak compared with the pre-outbreak period, accounting for temporal trends and variation in other risk factors. In the first stage, at province level, we applied a quasi-Poisson time series regression model with smooth spline functions for time variables and observed predictors.^15^ Specifically, we included a linear term for time for the long-term trend, a cyclic cubic B-spline with three equally-spaced knots for day of the year to model seasonality, and indicators for day of the week to take into account weekly variation in mortality. The excess risk in mortality during the COVID-19 outbreak was defined through a constrained quadratic B-spline with four equally-spaced knots for the days from 01/02/2020 to 15/05/2020 (outbreak period). This function constraints the excess risk to start from null at the beginning of February, and then allows it to vary flexibly until the end of the study period. Potential differences in the underlying mortality risk due to non-optimal weather between the pre-outbreak and outbreak periods where controlled by including a term for mean daily temperature. This was defined through a distributed lag non-linear model (DLNM) over 0-21 lag days,^16^ using a cross-basis parameterization defined in previous studies.^17^ It was decided not to control for influenza in the main model, as laboratory testing could be affected by the pandemic, thus resulting in artificial reduction in flu incidence. This choice was tested in a sensitivity analysis, where we controlled for the effect of influenza using a two-week moving average of the reconstructed daily incidence.

The province-specific coefficients of the function defining the excess risk were then pooled in the second stage using a mixed-effects multivariate meta-analysis,^18^ and best linear unbiased predictions for each of the 107 provinces were extracted. This approach allows borrowing strengths in the estimates across provinces, while at the same time modelling flexible associations of multi-parameter functions.^19^

### Quantification of excess mortality

The model was performed on the province-specific aggregated series composed by the 7,270 municipalities with full data, and the estimates were used to compute the relative risk (RR) representing the excess in each province for each day of the outbreak period. We then reconstructed the complete mortality series for all the municipalities by using the data in the period 2015-2019 to compute, in each province, the proportion of deaths occurring in municipalities with full data, and then applying the same proportion to the four months of 2020. The daily number of excess deaths was computed as *(RR-1)/RR*n*, where *n* is the daily number of deaths, and aggregated by week during the outbreak period (from the 1^st^ of February), and then in total for the period with reported COVID-19 mortality (from the 15^th^ of February). Excess number and fraction of deaths were then computed for each province, region, area (North, Central, South, and Islands) and for the whole Italy. We computed empirical confidence intervals (eCI) at 95% by Monte Carlo simulation of the coefficients of the quadratic B-spline modelling the excess risk, assuming a multivariate normal distribution using the point estimates and (co)variance matrix. Sex and age-specific models were run separately to compute the excess across sub-groups of the population.

Information on repositories, software, and code for accessing the data and replicating the analysis are provided in the Section Availability of data and materials below.

## Results

In the three months from 15^th^ of February to 15^th^ of May 2020, approximately 208,320 deaths were registered in Italy, with an estimated increase of 47,490 (95%eCI: 43,984 to 50,362) from the expected baseline number when accounting for temporal trends and differences in temperature distribution (Table 2). This figure corresponds to a percentage excess of 29.5% (95%eCI: 26.8 to 31.9%). As previously reported, the excess shows a strong geographical pattern, with an extremely high rise in the number of deaths in the northern regions. Lombardy alone experienced a staggering 25,782 excess deaths, and together with two other northern regions (Veneto and Emilia-Romagna) accounts for 71.0% of all the excess mortality estimated in Italy. The maps in Figure 1 provides a better representation of the geographical distribution, showing a percentage increase in deaths that reaches almost 400% in few selected northern provinces. The excess mortality is comparatively milder in Central Italy, and considerably smaller or absent in the South and the islands, although with a patchy distribution and isolated spots such as in the provinces of Pesaro-Urbino (East coast) and Crotone (the ‘bottom of the foot’ in the South).

**Table 2.** Number of total deaths and estimates excess (with 95% empirical confidence intervals) during the intense period of the COVID-19 pandemic (15^th^ of February-15^th^ May 2020) in Italy, by sex.

|  | Both sexes |  | Males |  | Females |  |
| --- | --- | --- | --- | --- | --- | --- |
|  | Total | Excess | Total | Excess | Total | Excess |
| **Piemonte** | 18,417 | 5,577 (5,095 to 6,004) | 8,860 | 2,756 (2,577 to 2,898) | 9,557 | 2,810 (2,543 to 3,046) |
| **Valle d'Aosta** | 541 | 194 (159 to 224) | 253 | 85 (65 to 103) | 289 | 111 (87 to 129) |
| **Lombardia** | 50,874 | 25,782 (24,806 to 26,703) | 25,178 | 13,320 (12,844 to 13,673) | 25,697 | 12,447 (12,098 to 12,731) |
| **Trentino-Alto Adige** | 3,688 | 1,227 (1,057 to 1,384) | 1,700 | 518 (494 to 533) | 1,987 | 709 (613 to 787) |
| **Veneto** | 14,905 | 2,317 (1,793 to 2,776) | 7,120 | 1,146 (985 to 1,263) | 7,785 | 1,185 (727 to 1,582) |
| **Friuli-Venezia Giulia** | 4,135 | 461 (299 to 601) | 1,947 | 243 (147 to 310) | 2,188 | 213 (105 to 308) |
| **Liguria** | 7,599 | 2,346 (2,072 to 2,595) | 3,570 | 1,099 (1,039 to 1,146) | 4,029 | 1,249 (1,168 to 1,309) |
| **Emilia-Romagna** | 18,249 | 5,605 (5,212 to 5,938) | 8,906 | 3,020 (2,858 to 3,122) | 9,343 | 2,573 (2,333 to 2,784) |
| **Toscana** | 12,210 | 1,416 (1,137 to 1,643) | 5,897 | 794 (493 to 1,037) | 6,313 | 633 (450 to 772) |
| **Umbria** | 2,651 | 83 (-51 to 200) | 1,233 | 15 (-81 to 104) | 1,418 | 80 (-22 to 170) |
| **Marche** | 5,776 | 1,415 (1,283 to 1,523) | 2,852 | 817 (769 to 842) | 2,924 | 612 (473 to 715) |
| **Lazio** | 13,972 | -548 (-1,032 to -97) | 6,786 | -154 (-421 to 65) | 7,187 | -325 (-646 to -33) |
| **Abruzzo** | 4,162 | 419 (181 to 644) | 2,062 | 212 (135 to 277) | 2,101 | 225 (99 to 322) |
| **Molise** | 931 | 23 (-42 to 80) | 442 | 3 (-56 to 48) | 489 | 31 (-15 to 66) |
| **Campania** | 13,816 | 125 (-334 to 524) | 6,937 | 257 (-176 to 655) | 6,879 | -98 (-377 to 132) |
| **Puglia** | 10,939 | 685 (414 to 885) | 5,406 | 363 (75 to 602) | 5,533 | 322 (66 to 527) |
| **Basilicata** | 1,715 | -6 (-126 to 86) | 805 | -28 (-100 to 25) | 911 | 34 (-47 to 104) |
| **Calabria** | 5,396 | 119 (-40 to 244) | 2,660 | 83 (-101 to 244) | 2,736 | 53 (-131 to 209) |
| **Sicilia** | 13,742 | 11 (-564 to 487) | 6,672 | 24 (-271 to 258) | 7,069 | 70 (-179 to 252) |
| **Sardegna** | 4,602 | 238 (53 to 407) | 2,282 | 82 (-68 to 210) | 2,320 | 190 (-9 to 344) |
| **Italy** | 208,320 | 47,490 (43,984 to 50,362) | 101,568 | 24,655 (22,604 to 26,215) | 106,754 | 23,125 (20,997 to 24,609) |

**Figure 1.**
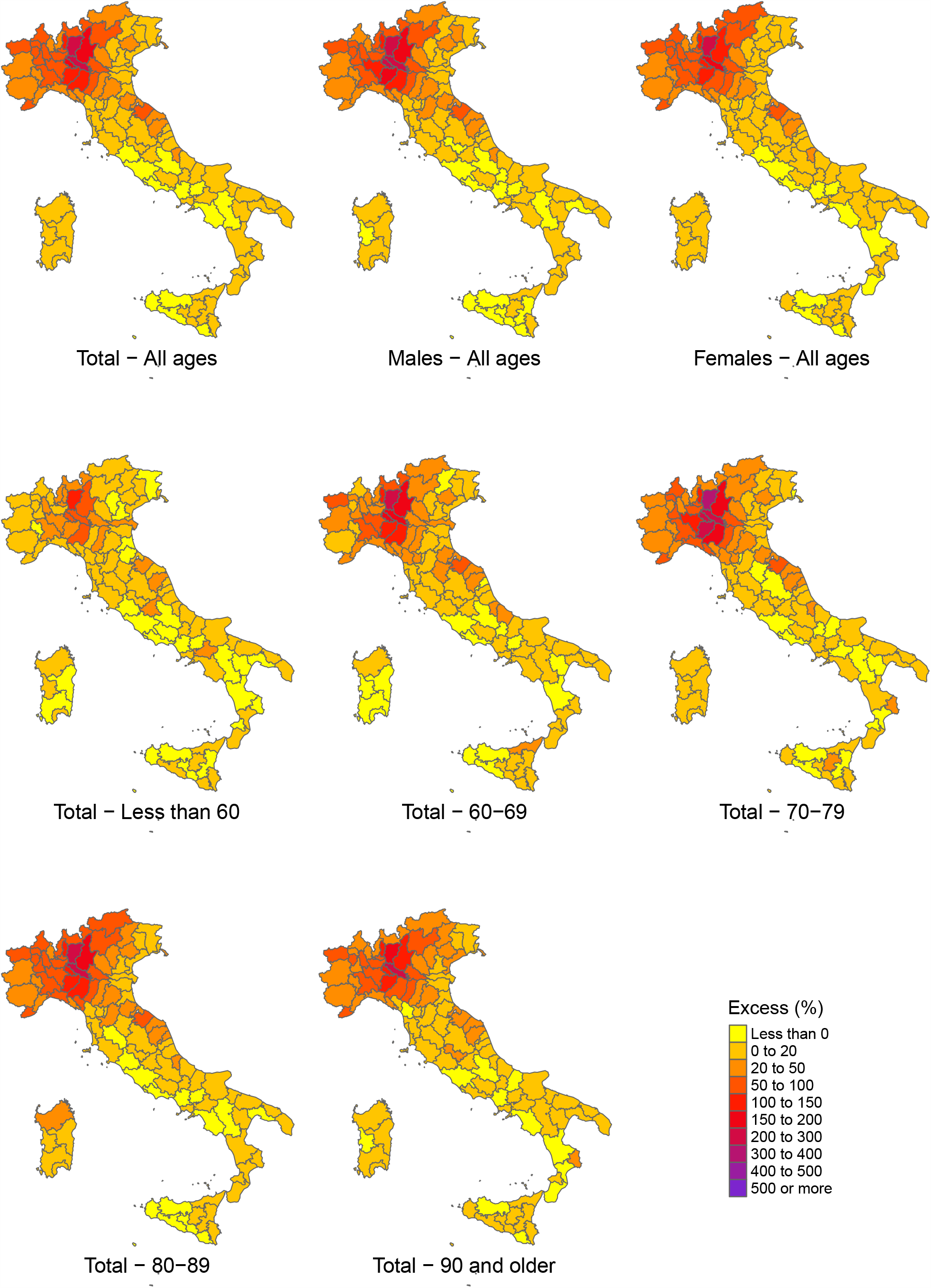
Maps of percentage excess in mortality during the intense period of the COVID-19 pandemic (15^th^ of February-15^th^ May 2020) by province in Italy, in total and stratified by sex and age group.

Differently from what reported during the outbreak, the increase in mortality is only slightly higher in men compared to women, with a total of 24,655 and 23,125, corresponding to percentage increases of 32.1% (95%eCI: 28.6 to 34.8%) and 27.7% (95%eCI: 24.5 to 30.0%), respectively (Table 2). Figure 2 reports the results stratified by age and sex, indicating a lower excess mortality in women in the age groups between 60 and 89, but almost identical to males in younger and older people. Excess mortality was substantially lower in people less than 60 years old, although this group still shows a percentage excess of 10.2% (95%eCI: -3.5 to 23.2%) in the outbreak period. The maps in Figure 1 suggests a similar geographical distribution of the excess deaths across sex and age groups. Figure 3 shows the trend in relative risk of mortality during the COVID-19 outbreak period by sex, age group, and area. The graphs suggest that the risk started increasing above the baseline at the beginning of March, and that the peak was reached at the end of the same month, almost 20 days after the implementation of lockdown measures. Given the uneven geographical distribution of the risk, with the densely populated provinces of the North mostly affected, this increased risks translates into a larger total excess in mortality, which reached 78.7% (95%eCI: 70.6 to 85.5%) above the baseline in the week of 18-24 March (figures reported in the Online Appendix). The risk then decreased during April, with evidence of an excess mortality until at least the first week of May. The temporal distribution of risk is similar across sex and age groups (top and middle panels), with an indication of a slight delayed peak for women and very elderly (90 years old and older). The relative risk for the less than 60 age group reaches a lower peak just above 1.2, and it returns to the baseline level by mid-April. Consistently with what discussed above, the analysis by geographical area (bottom panel) suggests a very high mortality risk in the North, that across the region is more than twice at the peak (RR above 2.0), while other areas show milder increases and earlier peaks. Some provinces showed staggering increases, with the percentage excess in Bergamo reaching 858.7% (95%eCI: 771.9 to 969.5%) in the week 18-24 March (see Online Appendix). There is some indication of an RR lower than 1 in the late stage of the epidemics in Central and Southern Italy, which can suggest an ‘harvesting effect’ with part of the deaths only anticipated by few weeks.^20^

**Figure 2.**
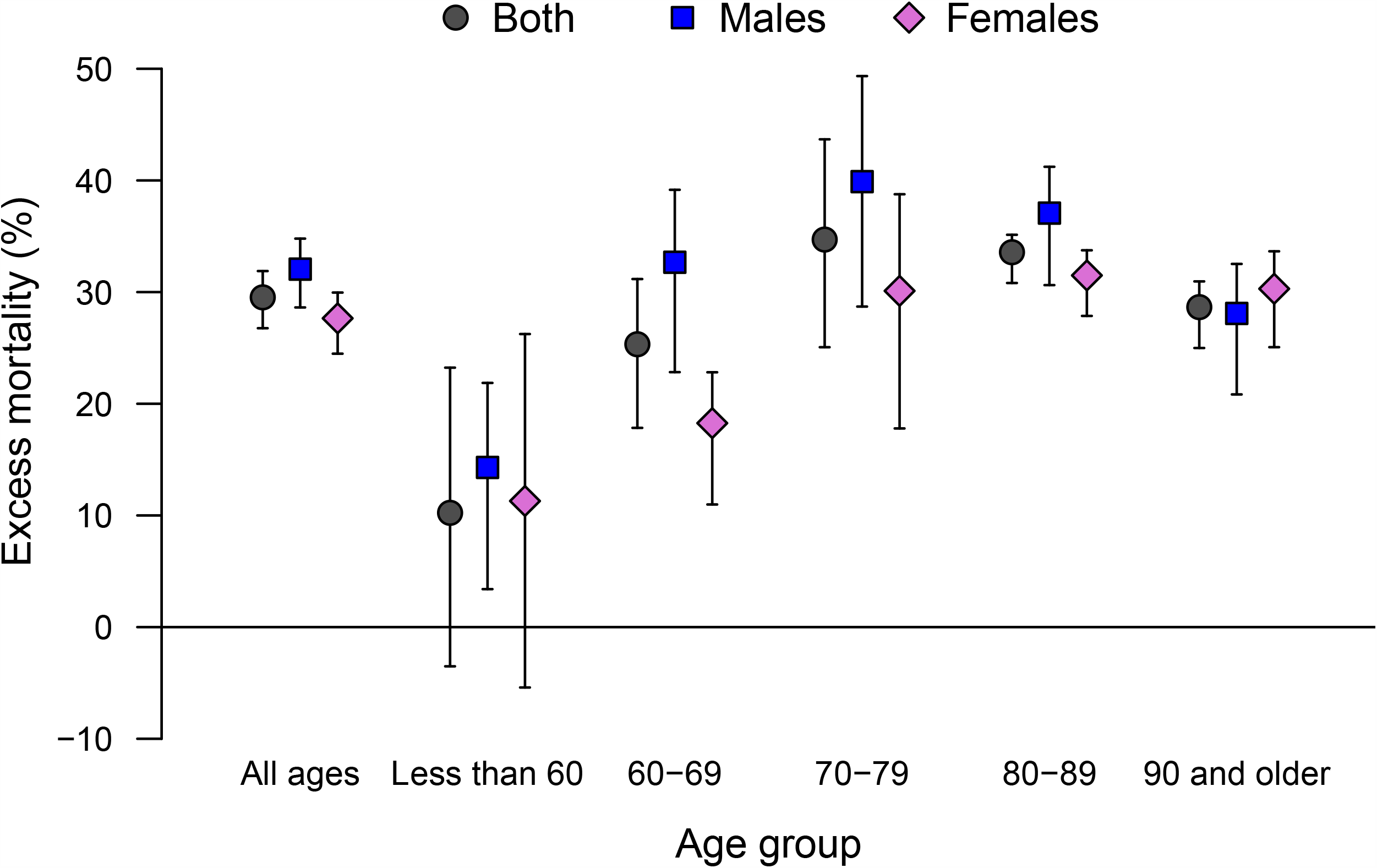
Percentage excess in mortality (with 95% empirical confidence intervals) during the intense period of the COVID-19 pandemic (15^th^ of February-15^th^ May 2020) in Italy, in total and stratified by sex and age group.

**Figure 3.**
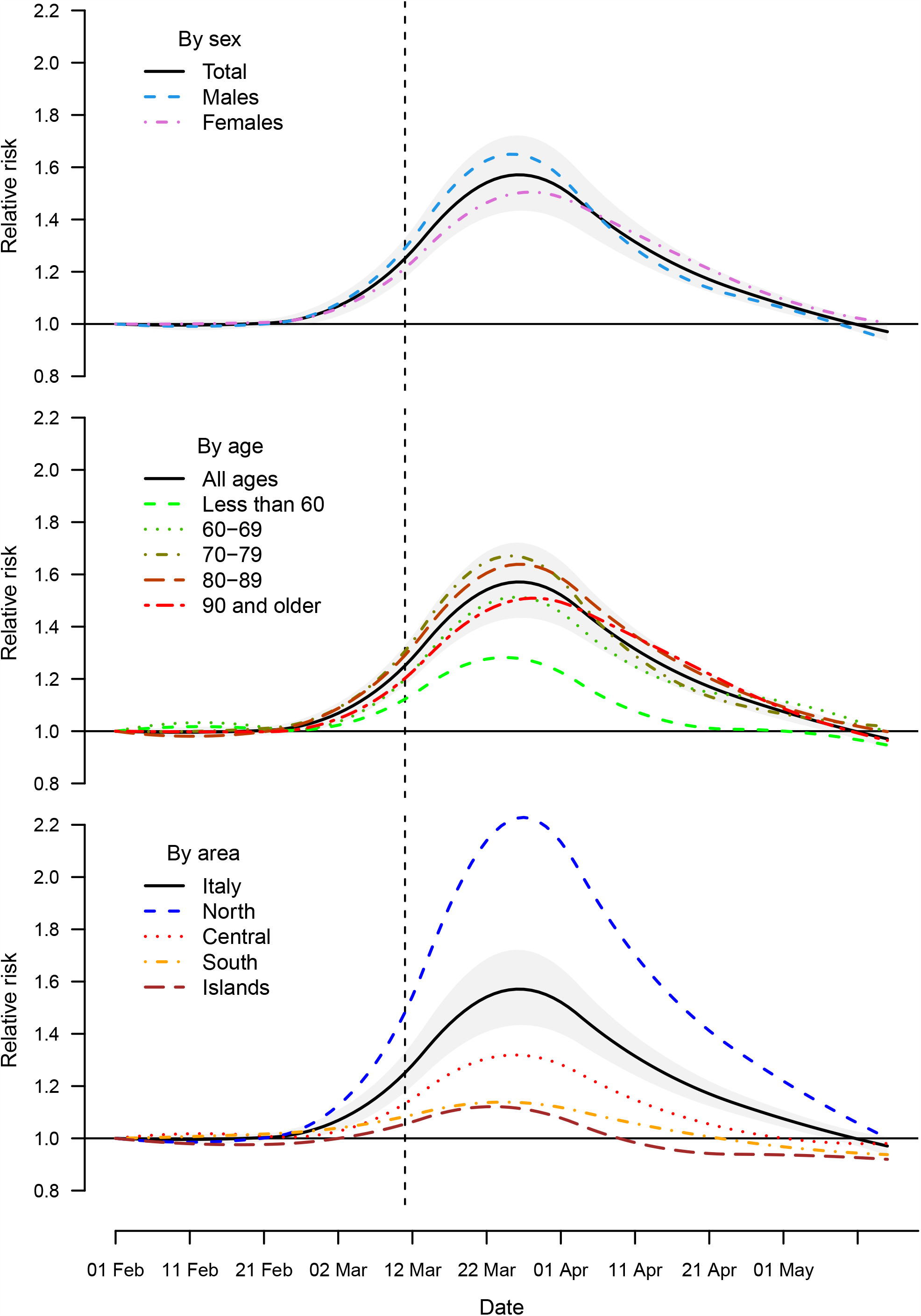
Trend in excess risk (relative risk, RR) during the period 1^st^ of February-15^th^ May 2020 in Italy by sex, age, and geographical area, compared with the total (with a band corresponding to the 95% empirical confidence intervals).

Controlling for the additional effect of influenza has little impact, with an estimated excess of 48,569 compared to 47,490 in the main model.

The full set of results, including number and fraction of excess deaths (with 95%eCI) by geographical aggregation (provinces, region, and full country), sex, age groups, and period (15^th^ of February - 15^th^ of May 2020, and then by week starting from the 1^st^ of February) is provided in the Online Appendix.

## Conclusions

This study offers a detailed assessment of mortality for all causes in Italy during the COVID-19 pandemic wave in the first months of 2020. The estimate of an excess of 47,490 deaths is far higher than the number of 31,610 deaths officially recorded as related to COVID-19 in the same months. The increased mortality risk shows a strong geographical pattern, with few northern provinces carrying the majority of excess deaths. Stratified analyses indicate differences in the impact by sex and age, while the analysis of the trend shows that excess mortality peaked the end of March. The most striking result of the analysis is the strong north-to-south geographical gradient in the impact of COVID-19 in Italy, consistent with previous studies from Italy.^8-10^ The vast majority of excess deaths occurred in selected northern provinces. However, differently from what previously reported,^10^ this study is able to identify significant increases in many provinces and regions of the South as well. The reasons for such north-south differences are still debated. A role was likely being played by the timing and characteristics of the SARS-CoV-2 outbreak, which started in provinces of the North with established trade connections with China and was amplified by the higher population density and internal travel.^21^ The implementation of strict lockdown policies, including the restriction in inter-regional movements, have likely contributed to reduce the spread of the infection and the consequent increased mortality in other regions, particularly in the South. This aspect can also explain the patchy map of risk across the country, where hotspots of risk can be the result of the influx of infections from high-prevalence provinces of the North before strict lockdown policies where implemented. Other suggested reasons of the geographical distribution of excess mortality are the occurrence of mass gathering events that boosted local numbers of infections,^22^ and regional differences in health service organization, socio-economic determinants, and urbanization.^23-25^

The assessment of results of stratified analysis offers interesting clues on sex and age differentials. While this study confirms the higher risk in men, the difference is comparatively small, with a percentage increase of 32.1% versus 27.7% in women. The analysis by age in Figure 2 reveals that the highest excess occurred among the 70-79 years old, but it is lower in younger and elderly groups. The risk is even lower but still present in the rest of the population, with evidence of excess deaths even in people under 60 years old. The graph in Figure 3 shows instead some differences in the timing of the excess mortality across sex, age groups, and area. The more delayed peak in women and very elderly can be indicative of differential risk pathways, for instance related to reduction in access to emergency services during the lockdown or the delayed surge of infections in nursing care homes. The early decrease and then below average mortality in Central and Southern Italy can instead imply that some of the excess is due to anticipation of death in frail individuals. These suggestive results can form the basis of future analyses.

The figures reported in this study can be compared with those previously published in peer-reviewed articles, although the comparison is made difficult by the different study population, study period, data collection procedures, and analytical methods. Michelozzi and colleagues analysed data collected within a surveillance system from 19 Italian cities, reporting a total excess mortality of 45% from the beginning of March until the 18^th^ of April 2020.^9^ The increase was higher in northern cities, in men, and in the elderly, although with estimates that are somewhat different from the present analysis, in part probably due to the focus to urban areas and the different period. Magnani and colleagues used data released by ISTAT on 4,433 municipalities covering the whole Italian territory, and quantified the extrapolated excess to the total Italian population in 44,352.5 and 680.4 deaths in persons older and younger than 60, respectively, between the 1^st^ of March and the 15^th^ of April.^10^ They however reported negative estimates in several regions, in particular for the younger group. These differences can be due to the simple pre-post comparison adopted in these studies, which does not account for temporal trends and for the milder weather conditions in 2020.

This study benefits from the application of advanced two-stage design and statistical models that allows a flexible estimation of the excess mortality risk and a definition of baseline risk that considers temporal trends and variations in known risk factors. The analysis is performed at province level and stratified by sex, age, and week periods, thus offering a fine characterization of the impact of the COVID-19 pandemic in Italy. Data and code are available, making the analysis entirely reproducible, and the full set of results is released in the appendix and through a web application. Some limitations must however be acknowledged. First, the low number of cases prevents the full application of the two-stage modelling in age groups less than 50 years old, and therefore to address the question about to which extent COVID-19 affects young people. Furthermore, the interrupted time series design defines a simple pre-post comparison that cannot inform about the various mechanisms leading to increased risks. The question about controlling or not for incidence of seasonal influenza is emblematic in respect to this problem. Similarly, while the analysis of all-cause mortality can provide a good estimate of the overall burden, it offers no information about the various mechanisms, patho-physiological or of different nature, leading to increased deaths. Finally, the analysis is based on an early-release dataset from ISTAT that has not been consolidated, and this may lead to an underestimation of the number of deaths that occurred in 2020 and therefore of the excess during the pandemic. These issues can be addressed when additional data and information on the COVID-19 pandemic will be released.

In conclusion, this study offers a detailed picture of excess during the first months of the COVID-19 pandemic in Italy. The analysis indicates that excess mortality during the outbreak has been far higher than the number of deaths officially reported as related to COVID-19, with strong geographical differences and patterns of risk that varies by sex, age, and period. The figures can be used in future research to explain differential risks across regions and sub-groups, and the role of national or local policies implemented during the period.

## Data Availability

The data, software and code for replicating the analysis and full set of results are made available. The analysis was performed in the R software environment. The scripts for downloading the data and for fully replicating the analysis and results are available in a GitHub repository (https://github.com/gasparrini/ItalyCOVIDdeath). The data (release on 18/06/2020 with updated mortality up to 15th of May) are extracted from the ISTAT website (https://www.istat.it/it/archivio/240401) and stored in the repository. A web application is made available at (https://mscortichini.shinyapps.io/app20200703/) to visualize maps of the excess risk and extract the full set of results, which are also available in the Online Appendix of this article.

https://github.com/gasparrini/ItalyCOVIDdeath

https://mscortichini.shinyapps.io/app20200703/

## Funding

This work was supported by the Medical Research Council-UK (Grant ID: MR/M022625/1) and the European Union’s Horizon 2020 Project Exhaustion (Grant ID: 820655).

## Data Availability Statement

The data, software and code for replicating the analysis and full set of results are made available. The analysis was performed in the R software environment. The scripts for downloading the data and for fully replicating the analysis and results are available in a GitHub repository (https://github.com/gasparrini/ItalyCOVIDdeath). The data (release on 18/06/2020 with updated mortality up to 15^th^ of May) are extracted from the ISTAT website (https://www.istat.it/it/archivio/240401) and stored in the repository. A web application is made available at (https://mscortichini.shinyapps.io/app20200703/) to visualize maps of the excess risk and extract the full set of results, which are also available in the Online Appendix of this article.

## Acknowledgements

This study was based on finely stratified mortality data made publicly available by ISTAT in an early-release database published on 18/06/2020.

## Conflict of interests statement

The authors declare that they have no competing interests.

## Notes

### Competing Interest Statement

The authors have declared no competing interest.

